# The association between COVID-19 and preterm delivery: A cohort study with a multivariate analysis

**DOI:** 10.1101/2020.09.05.20188458

**Authors:** Spanish Obstetric Emergency Group, Oscar Martínez Pérez

**Affiliations:** Encompassing 45 hospitals; Department of Obstetrics and Gynaecology. Hospital Universitario Puerta de Hierro. Majadahonda, Madrid, Spain

## Abstract

**Objective:** To determine whether severe acute respiratory syndrome coronavirus 2 (SARS-CoV-2, the cause of COVID-19 disease) exposure in pregnancy, compared to non-exposure, is associated with infection-related obstetric morbidity.

**Design and setting:** Throughout Spain, 45 hospitals took part in universal screening of pregnant women going into labour using polymerase-chain-reaction (PCR) for COVID-19 since late March 2020.

**Methods:** The cohort of exposed and unexposed pregnancies was followed up until 6-weeks post-partum. Multivariate logistic regression analysis, adjusting for known confounding variables, determined the adjusted odds ratio (aOR) with 95% confidence intervals (95% CI) of the association of COVID-19 exposure, compared to non-exposure, with infection-related obstetric outcomes.

**Main outcome measures:** Preterm delivery (primary), premature rupture of membranes and neonatal intensive care unit admissions.

**Results:** In the cohort of 1,009 screened pregnancies, 246 were COVID-19 positive. Compared to non-exposure, COVID-19 exposure increased the odds of preterm birth (34 vs 51, 13.8% vs 6.7%, aOR 2.12, 95% CI 1.32–3.36, p = 0.002), premature rupture of membranes at term (39 vs 75, 15.8% vs 9.8%, aOR 1.70, 95% CI 1.11–2.57, p = 0.013) and neonatal intensive care unit admissions (23 vs 18, 9.3% vs 2.4%, aOR 4.62, 95% CI 2.43 – 8.94, p< 0.001).

**Conclusion:** This first prospective cohort study demonstrated that pregnant women infected with SARS-CoV-2 have more infection-related obstetric morbidity. This hypothesis merits evaluation of a causal association in further research.

## Introduction

Severe acute respiratory syndrome coronavirus 2 (SARS-CoV-2), identified in December 2019, is the cause of the illness named COVID-19.^1,2^ With more than 249,000 confirmed cases and more than 28,700 deaths by 20^th^ August 2020, Spain remains one of the European countries most severely affected by the ongoing COVID-19 pandemic.^3,4^ Spain also established a universal screening programme for pregnancies in light of the higher disease exposure. We observed that obstetric intervention may influence the clinical course of the disease.^5^ The cohort of pregnant women assembled through this programme lends itself to evaluation of concerns about obstetric outcomes.

The majority of non-pregnant patients with COVID-19 have uncomplicated or mild illness (81%), some will develop severe illness associated with cytokine-mediated inflammation phenomena such as IL-6 associated with the need for mechanical ventilation.^6^ Initial studies have reported similar involvement in pregnant patients.^7^ The inflammatory mediators associated COVID-19 have previously been related to poor perinatal outcomes.^8–9^ This background naturally leads to the question as to whether COVID-19 affects pregnancy adversely.

We hypothesised that COVID-19 exposure in pregnancy, compared to non-exposure, would increase infection-related obstetric morbidity including preterm birth and premature rupture of membranes which in turn would increase the admissions of the neonate to intensive care units. We tested the hypothesis in a multivariate logistic regression analysis, adjusting for the effect of known confounding variables.

## Materials and methods

In March 2020, the Puerta de Hierro University Hospital in collaboration with 76 hospitals of several Spanish regions, launched OBS COVID REGISTRY, a population-based, longitudinal observational and analytic study, to quantify the obstetrical and perinatal morbimortality possibly associated to COVID infection throughout Spain.^10^ Between the third week of March and the first week of May 2020, 45 hospitals started consecutive universal PCR screening of all patients going into labour (Figure 1).

**Figure 1:**
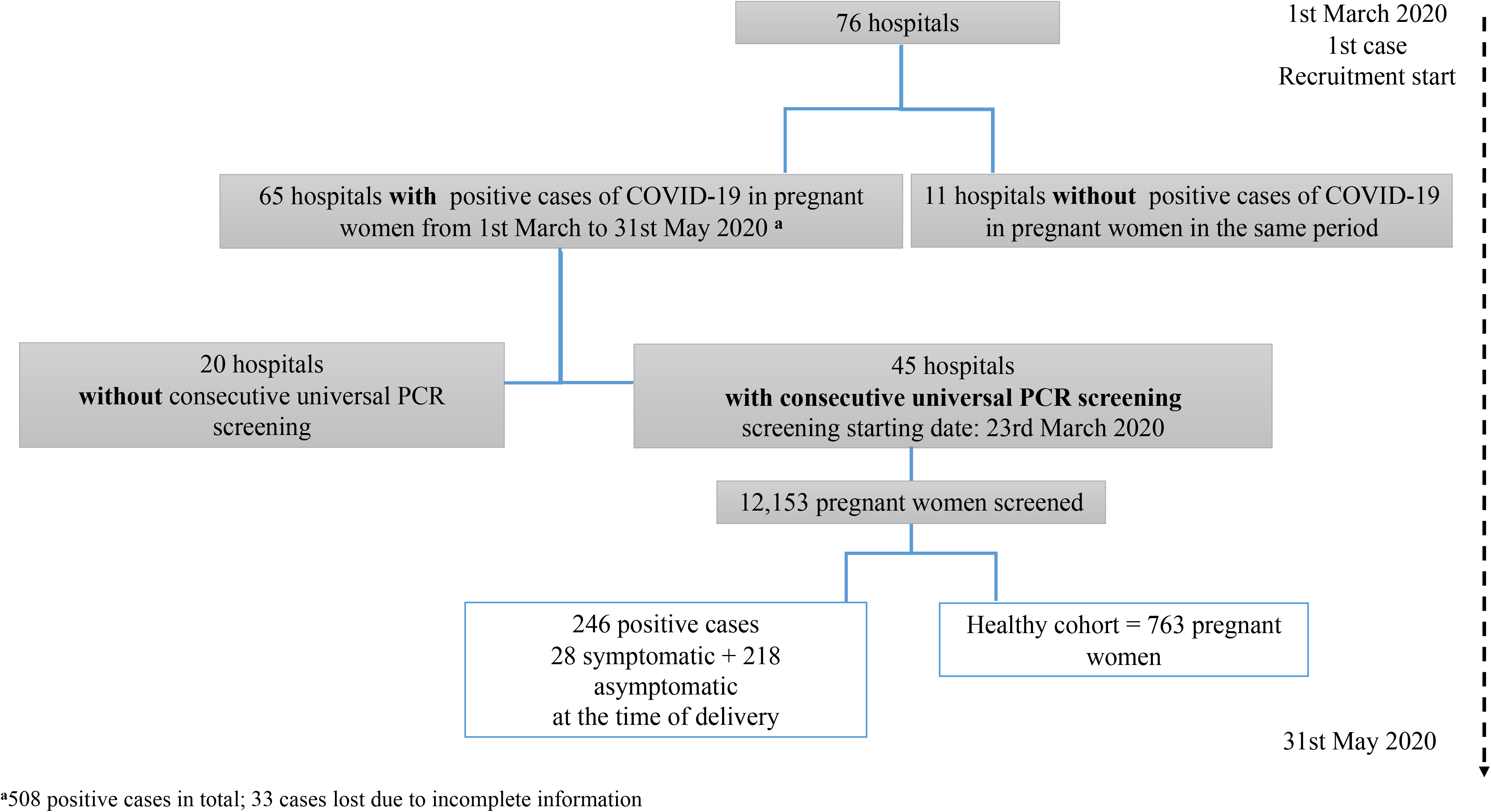
Study Flow chart

### Ethics Committee Approval

All procedures were approved by Puerta de Hierro University Hospital (Madrid, Spain) ethics committees (reference number, 55/20). All the 76 hospitals involved in the previously created Emergency Obstetrics Spain Group recognized and adopted the procedures. Written informed consent was obtained, sometimes in a delayed fashion, from the study patients at each centre when possible. The majority of the informed consents were provided verbally depending on local ethics committee regulations and special policies issued for Covid-19 research due to the pandemic situation. All verbal consents were recorded in the patient medical records. The registry protocol, including all the information on the variables under study, is available in the OSF website: https://osf.io/k82ja/?view_only?61d3773cc12f423b8568ff11acb0fa44We developed an analysis plan using recommended contemporaneous methods and followed existing guidelines for reporting.^11^

### Assembly of the cohort

Case recruitment for this initial study ended on 31^st^ May 2020. Hospitals collected the encoded information in two separate phases: during the enrolment period that occurred at the time of the SARS-CoV-2 test during pregnancy and within 2 weeks after birth. Case eligibility criteria were pregnant patients with laboratory confirmation of COVID-19 infection, irrespective of clinical signs and symptoms or the result of another serological test; positive RNA polymerase-chain-reaction (PCR) test from nasopharyngeal and/or oropharyngeal swabs from mother. In those cases, with a clinical presentation of COVID19, it was classified following the WHO division for adults: mild symptoms, mild-moderate pneumonia, severe pneumonia and septic shock.^12^ All identified cases were included in the study. The healthy cohort (comparison group not exposed to COVID-19) included pregnant women who were negative for the PCR test, who were selected in each center based on the delivery date of the cases, selecting deliveries before and/or after that of the case,with maximum 3 healthy deliveries per each infected pregnant woman (Figure 1). SARS CoV2 negative patient information collection used the same templates and same database as SARS CoV2 positive. The list of hospitals included in the study with their corresponding reported cases is in Appendix 1.

Information regarding the demographic characteristics of each pregnant woman, comorbidities and current obstetric history was extracted from the clinical history and from the interview with the patient; subsequently, age and race were categorized following the classification used by the CDC.^13^ Definitions of obstetric conditions followed international criteria.^14–16^ Perinatal events, medical and obstetric complications were recorded. Patients were followed until six weeks postpartum. Neonatal events were recorded until 14 days postpartum. A total of 33 dropout cases were recorded from the beginning of the registry; the reasons were incomplete information in the registry database, did not participate in the six-week postpartum follow-up and/or voluntary withdrawal of the patient.

### Data analysis

For the descriptive analysis of the data, absolute and relative frequencies were used in the case of categorical variables and means and ranges in the case of quantitative variables. The possible association of both the characteristics of the patients and the outcomes collected with COVID infection was analysed using the Pearson’s Chi-square test or Fisher’s exact test and the Mann-Whitney U test (after checking the absence normality of the data using the Kolmogorov-Smirnov test). Statistical tests were two-sided and were performed with SPSS V.20 (IBM Inc., Chicago, Il, USA); statistically significant associations were considered to exist when the p value was less than 0.05.

For computing measures of association of the outcomes of interest that were statistically significant in the univariate analysis (and with enough number of events) with COVID-19 infection, the influence of known and suspected measured confounding factors was controlled for multivariate logistic regression modelling in order to derive adjusted odds ratios (aOR) with 95% confidence intervals (95% CI). Models were built for each outcome separately, incorporating a range of independent variables appropriate for the adjustment of the association between COVID-19 infection and that outcome. The selection process for variables was driven by causal knowledge for the adjustment of confounding, based on previous findings and clinical constraints.^9,12–16^ Besides COVID positivity, the preterm delivery model included Ethnicity [categorized as white European, Latin American and other ethnic groups (black non-Hispanic, Asian non-Hispanic and Arab)], multiple pregnancy, in vitro fertilization, gestational hypertensive disorders (moderate or severe preeclampsia and HELLP), miscarriage risk and clinical and ultrasound prematurity screening; the spontaneous preterm delivery model included ethnicity (categorized as above), multiple pregnancy, miscarriage risk and clinical and ultrasound prematurity screening; the premature rupture of membranes at term (PROM) model included multiple pregnancy, miscarriage risk, cough, obesity (BMI>30 kg/m^2^) and smoking [categorized as smokers (actual and ex-smokers) and non-smokers]; the preterm premature rupture of membranes (PPROM) model included multiple pregnancy and miscarriage risk; and the neonate intensive care unit (NICU) admission model included multiple pregnancy, gestational hypertensive disorders and clinical and ultrasound prematurity screening as independent variables.

A complete list of the final set of covariates is provided with each model in the results section. The modelling was conducted after excluding cases with missing data. A prematurity screening program was not established in all participating hospitals and that variable had 11.3% of missing values, whereas the remaining variables had less than 1.2% of missing values. Regression analyses were carried out using lme4 package in R, version 3.4 (RCoreTeam, 2017).^17^

## Results

One thousand and nine (1,009) patients who gave birth between 23^rd^ March and 31^st^ May were analysed: 246 cases and a healthy cohort of 763 pregnant women who were negative for the PCR test. Of the 246 positive cases, 88.6% (n = 218) were asymptomatic at delivery while 11.4% (n = 28) were symptomatic. Of the asymptomatic women, 44 (20.2%) had previously presented symptoms and 174 (79.8%) were totally asymptomatic. On the other hand, of the pregnant women who showed symptoms at the time of delivery, 24 (85.7%) cases corresponded to mild symptoms (being the most prevalent, cough 33.3%, and anosmia 20.8%, followed by fatigue/discomfort, fever and dyspnoea), 2 (7.1%) pregnant women presented mild-moderate pneumonia and another 2 (7.1%) pregnant women had developed severe pneumonia. No case of septic shock or maternal death was recorded in pregnant women with COVID-19 included in the study.

The demographic characteristics, comorbidities and current obstetric history of the positive cases and the healthy cohort (246 vs 763) are shown in Table 1. The only variable in which statistically significant differences were observed was ethnicity, being significantly higher the proportion of Latin American women in the COVID-19 group (p < 0.001; OR = 2.85, 95% CI: 1.96–4.15), while the opposite was true for White European patients (p < 0.001, OR = 0.49, 95% CI: 0.36–0.67).

**Table 1:**
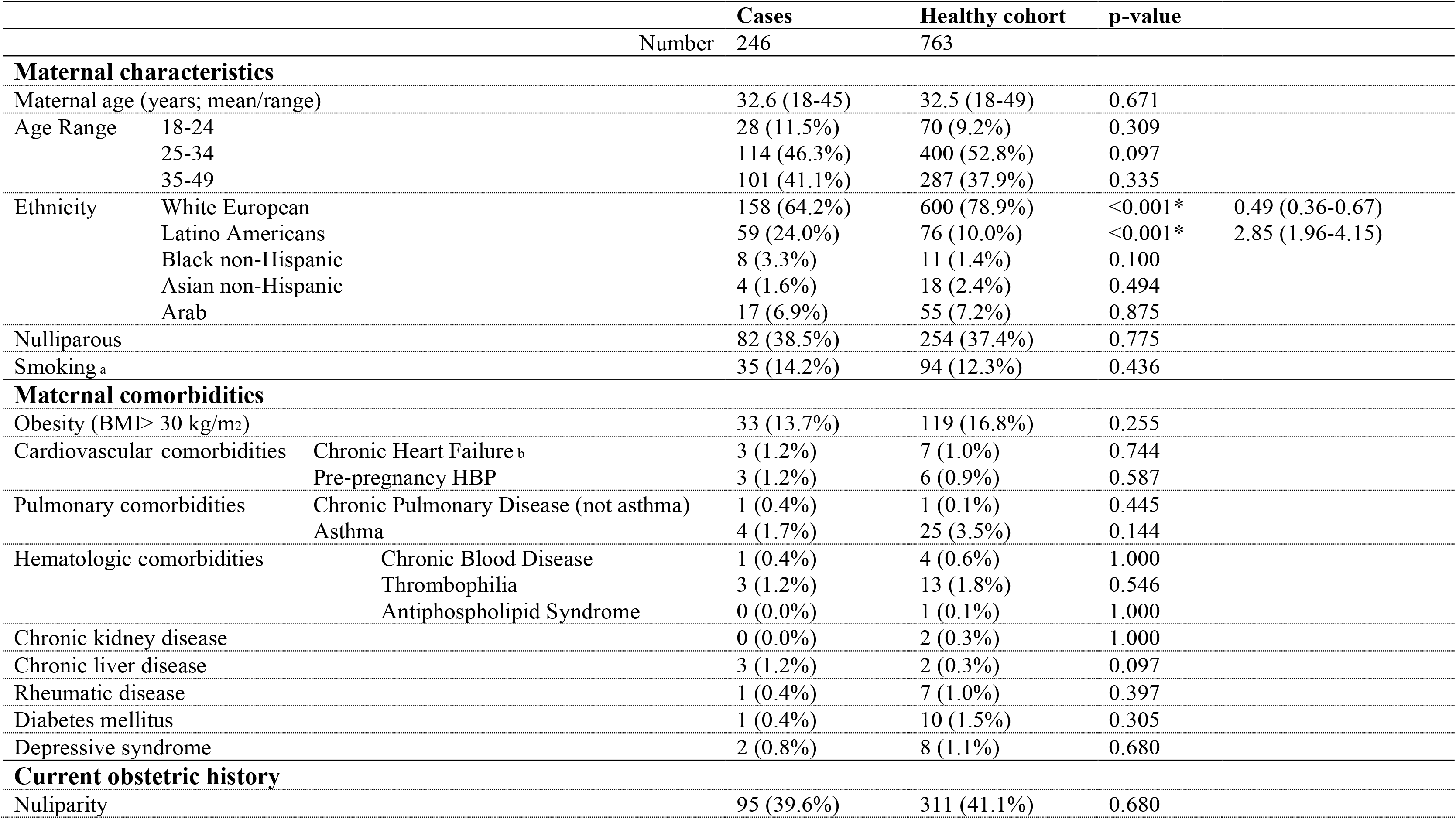
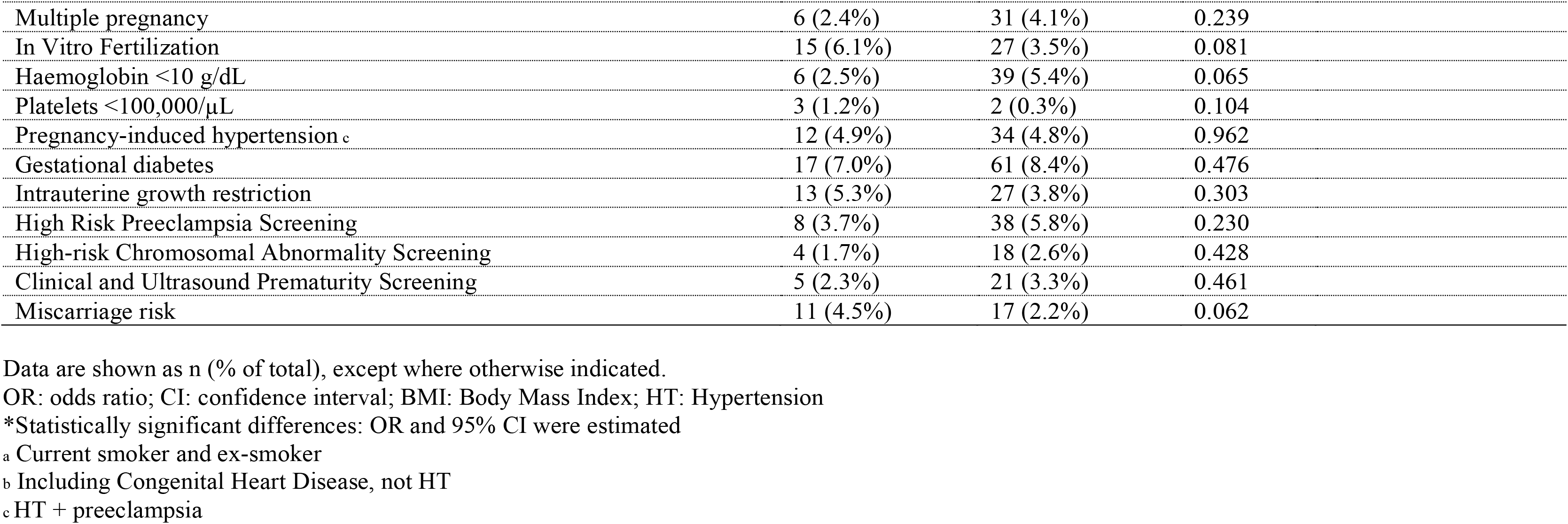
Patient demographic characteristics, comorbidities and current obstetric history in a pregnancy cohort screened for COVID-19.

When the possible association of perinatal and neonatal events with COVID-19 was analysed by univariate and multivariate logistic regression using complete case analyses (without imputation for missing values) (Table 2 and 3), twice as many deliveries with less than 37 weeks of gestation were observed among the cases (13.8%) than in the healthy group (6.7%) (p = 0.002), with an adjusted OR equal to 2.12 (95% CI: 1.32–3.36), although no statistically significant differences had been observed in the clinical and ultrasound screening for prematurity between both groups (p = 0.461) (Table 1). Among preterm deliveries, iatrogenic preterm delivery (not associated with PPROM) was practically four times more frequent in infected pregnant women than in non-exposed (healthy) pregnant women (4.9% vs 1.3%, p = 0.001), while the occurrence of spontaneous preterm deliveries was not affected by COVID-19 status (p = 0.760, adjusted OR = 1.10, 95% CI: 0.57–2.06) (Table 3). In the exposed group, symptomatic COVID-19 were present in 5 (42%) out of 12 iatrogenic preterm deliveries, while this was the case in only 3 (20%) out of 15 spontaneous preterm deliveries.

**Table 2:**
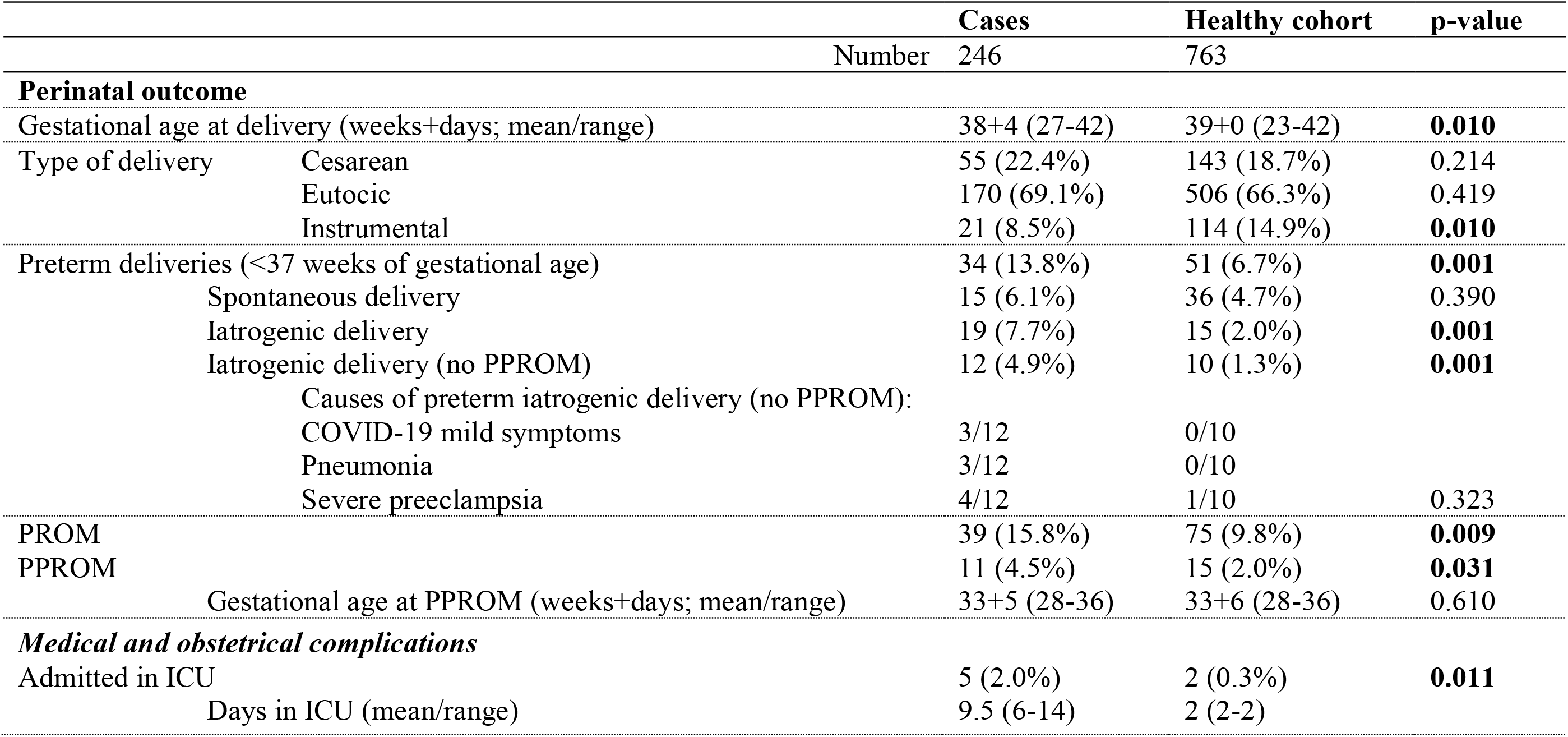
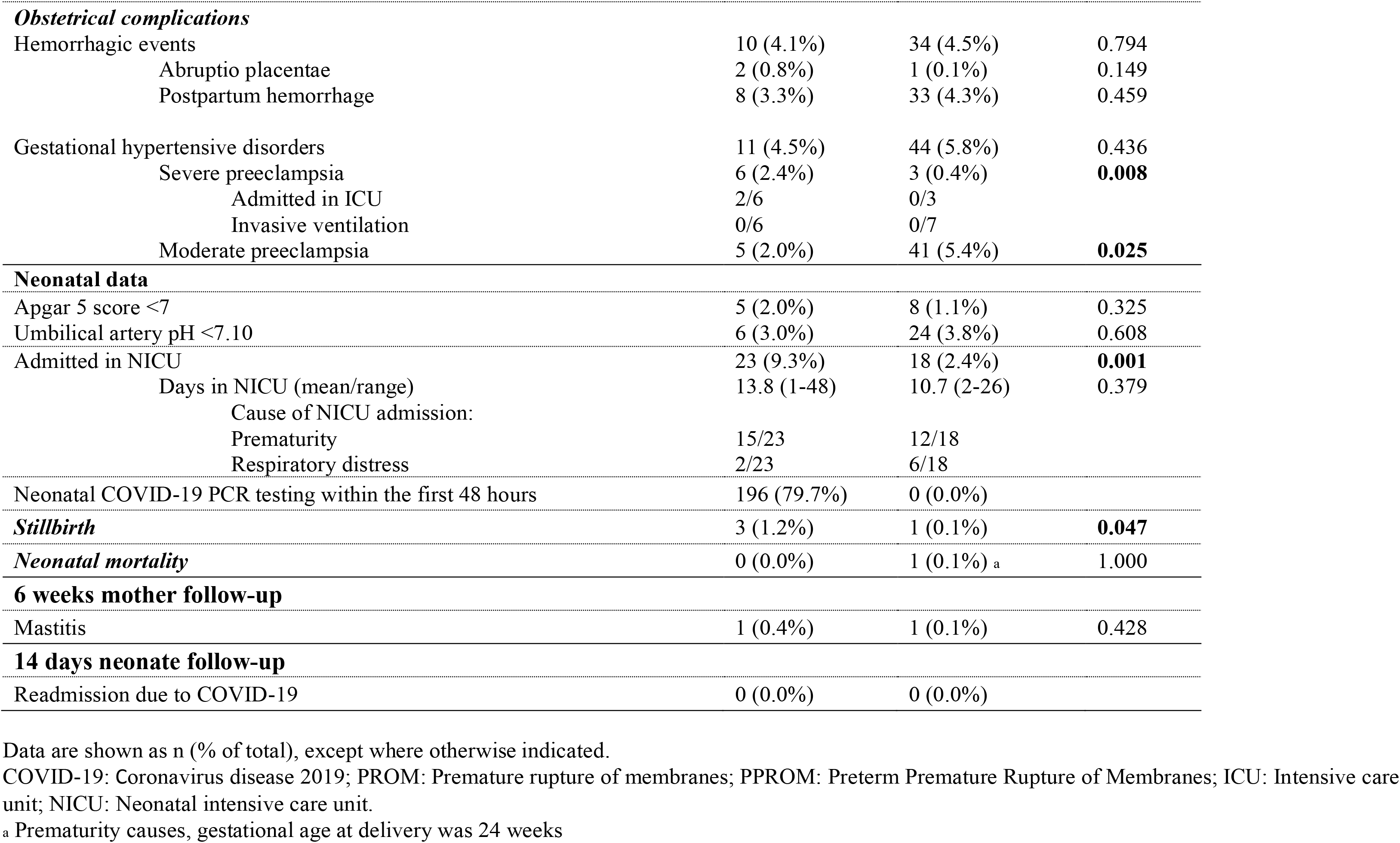
Maternal and neonatal outcomes in pregnant women screened for COVID-19.

**Table 3:**
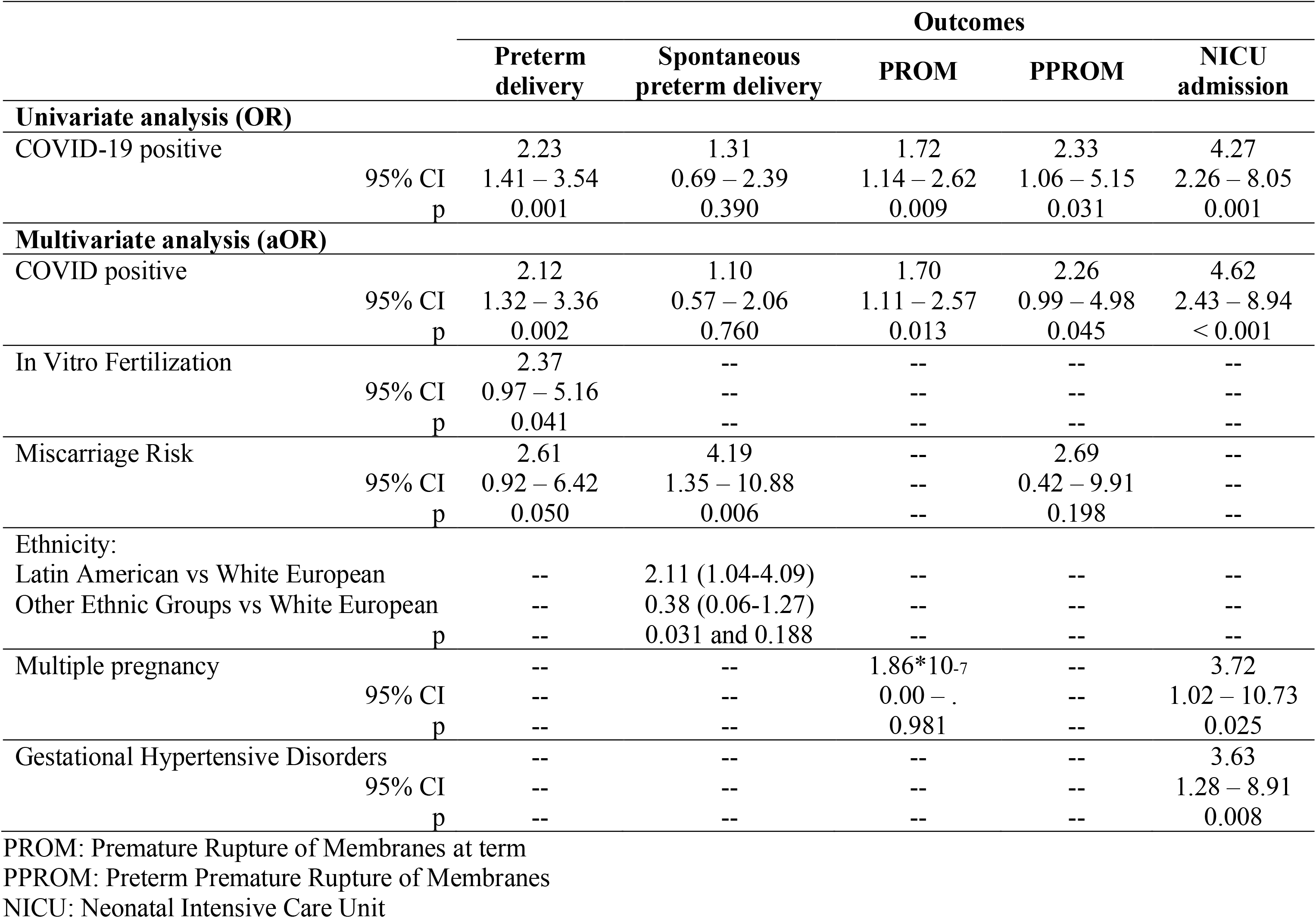
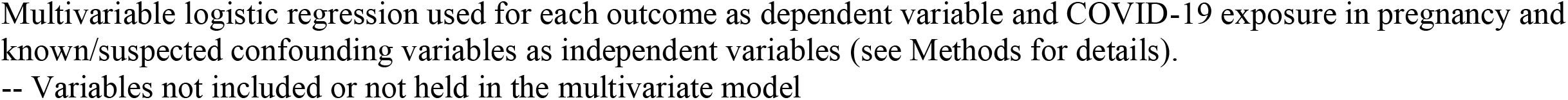
Odds ratio (OR) and adjusted odds ratio (aOR) with the corresponding 95% confidence intervals and p-values for the obstetric outcomes associated with exposure to COVID-19 in pregnancy.

Similarly, a higher risk of premature rupture of membranes, at term (PROM) and preterm (PPROM), was observed in the exposed group (p = 0.009 and p = 0.031, respectively) (Table 2). In the case of PROM, the finding of multivariate logistic regression was consistent with the above result, with a 70% increase of occurrence in the COVID-19 group compared to the non-exposed group (adjusted OR = 1.70, 95% CI:1.11–2.57) (Table 3).

No maternal deaths were recorded in the 1,009 patients in the study, but there were intrauterine foetal deaths, with the proportion of these being considerably higher in patients with COVID-19 than in the non-exposed ones (1.2% vs 0.1%, p = 0.047) (Table 2).

When the information regarding the neonate was analysed (Table 2 and 3), those born to mothers with COVID-19 were admitted to the NICU significantly more often than those born to healthy mothers (p < 0.001, adjusted OR = 4.62, 95% CI: 2.43 – 8.94). Prematurity and respiratory distress were the main causes of NICU admission (Table 2), while none of these admissions were due to COVID-19 disease in newborns. In 189 (76.8%) of the COVID-19 cases, a PCR analysis was performed on nasopharyngeal and/or oropharyngeal samples of the newborns; 147 were performed during the first 12 hours of life, three of which were positive, and another 42 were performed until 48 hours of life, all resulting negative. The 3-initial positive newborns were retested at 48 hours, with final negative results.

## Discussion

### Main findings

To our knowledge, this is the first prospective cohort study to analyse whether a relationship exists between COVID-19 exposure and infection-related obstetric outcomes. We found, using multivariate models adjusting for confounding factors, that the pregnant women with COVID-19 had more preterm births, premature rupture of membranes at term and NICU admissions compared to the pregnant woman who were not exposed.

### Strengths and weaknesses

Ours is a cohort study carried out during a difficult pandemic situation whose continuing objective is to investigate the influence of COVID-19 on delivery and the puerperium. We wish to obtain the best epidemiological information in the shortest possible time with a follow-up 6 weeks after delivery. Patient recruitment continues in our registry and this is an initial analysis. Our work is one of the first prospective cohort studies to analyse the relationship between COVID-19 and prematurity. The relationship that we establish with premature rupture of membranes raises future lines of research.

The most important limitation of our work is the inability to compare infected patients with healthy patients from the beginning due to the lack of diagnostic tests and the health sector crisis that occurred. When a screening system was established, there were not as many patients with severe symptoms and the number of events reduced the ability to analyse some effects of symptomatic COVID-19. Many cases of obstetric severe preeclampsia, haemorrhage, pulmonary thromboembolism and abruptio occurred mainly in the months of March and April before many centres started screening programmes and the cohort study began, so no distinction has been made between the different clinical presentations of the disease. We could not do a multivariate analysis of such conditions.

No serological test was performed on patients who had a negative PCR test, either because the tests were not available at the time of recruitment or because they did not have a proven sensitivity. In some cases, these patients may have already had the disease. No serology was performed during those months on asymptomatic PCR-positive patients to confirm their disease and immune response. Our study is best understood if the results are interpreted under this premise and therefore our group continues to recruit patients to seek more associations, explanations and causations. This work reflects the conditions of patients with COVID-19 at the time of delivery and the puerperium. It has not analysed the course of the disease during pregnancy, nor has it recorded late abortions, vertical transmission, or causes of intrauterine mortality.

### Comparison with other studies

The symptoms of the patients in our study do not differ from those already published.^18,19^ Although most did not have any symptoms, we did find an increase in obstetric pathology in these patients, which in our opinion indicates that in the pregnant woman with asymptomatic COVID-19 there is a specific obstetric pathology that needs to be recognised. In the same way as other authors, we have also found a demographic factor, such as ethnicity, that increases the possibility that a patient has COVID-19.^13,20^ It is necessary to know if there is a component of genetic susceptibility or if there are social factors that explain this association. There are already studies that relate this situation to less access to healthcare resources or the possibility of confinement which complies with healthcare measures.^21^

Patients with COVID-19 are at increased risk of preterm delivery associated with increased iatrogenic preterm delivery. The explanation for this risk is the need to end the pregnancy due to maternal diseases, such as severe pre-eclampsia and pneumonia, which are more frequent in these patients and lead to more labour inductions. A unique and novel finding in our study is the association between premature rupture of membranes at term and COVID-19. PROM may result in immediate risks and subsequent problems including maternal or neonatal infection.^22^ One of the possible explanations we found for this association is the activation of a series of mediators and biochemical pathways of inflammation in the premature rupture of membranes and premature delivery that are also found in COVID-19, such as macrophages or IL-6.^23^

The studies demonstrating the influence of IL-6 on preterm delivery are a strong basis for studying this association.^24^ Cytokines are vital in regulating immunological and inflammatory responses. Among them, IL-6 is of major importance because there is evidence that circulating IL-6 levels are closely linked to the severity of the COVID-19 infection.^25^ There are already treatments that are indicated based on these findings.^26^

We observed a significant increase in the stillbirth rate in the univariate analysis alone. The role of inflammation mediators in these deaths could be the subject of a line of research because it is known that women without COVID-19 who have a pregnancy loss, have significantly higher amniotic fluid IL-6 concentration levels than those with a normal outcome.^27^ We found no differences in mortality or early or late neonatal morbidity related to COVID-19 between healthy and infected patients in our series, unlike reports from other series.^28,29^ There is a higher risk that the children of COVID-19 mothers enter the NICU, with prematurity being one of the determining factors. All newborns were followed for at least 14 days by the different neonatology units of the participating hospitals, without any case of neonatal COVID-19 being detected in that period.

To date, there has been indirect evidence on placental involvement which would explain our findings.^30,31^ Our results derived using multivariate analyses confirm those of the cases series published at the beginning of the pandemic that described preterm deliveries and premature rupture of the membranes.^18,28,32^

## Conclusion

Pregnant COVID-19 patients are a population at risk of suffering preterm deliveries, and the disease has an impact on NICU admissions. Premature rupture of membranes at term and preterm are more frequent in patients with COVID-19.

## Data Availability

All variables of the data base  and  logistic regression strategies are available in OSF 
STROBE checklist for cohort studies have been  completed

https://osf.io/k82ja/?view_only=61d3773cc12f423b8568ff11acb0fa44

## Acknowledgements

Mr Jose Montes EFFICE.

Professor Khalid Khan for his scientific advice.

This study was fully funded with public funds obtained in competitive calls: grant COV20/00020 from the Institute of Health Carlos III and co-financed with FEDER funds.

